# The premature mortality to incidence ratio (PREMIER): a person-centred measure of cancer burden

**DOI:** 10.1101/2021.10.17.21265122

**Authors:** David Banham, Jonathan Karnon, Alex Brown, David Roder, John Lynch

**Affiliations:** School of Public Health, University of Adelaide, North Terrace, Adelaide, South Australia, 5000, Australia; Wardliparingga Aboriginal Health Research, South Australian Health and Medical Research Institute, North Terrace, Adelaide, South Australia, 5000, Australia; College of Medicine and Public Health, Flinders University, Sturt Road, Bedford Park, South Australia, 5042; Faculty of Health and Medical Sciences, University of Adelaide, North Terrace, Adelaide, South Australia, 5000; School of Health Sciences, Cancer Research Institute, University of South Australia, North Terrace, Adelaide, South Australia, 5000

**Author notes:** Corresponding author at: The South Australian Health and Medical Research Institute, North Terrace, Adelaide, South Australia, 5000.

**Keywords:** Indigenous Australians, cancer, premature mortality, mortality to incidence ratio, disparity

## Abstract

**Background:** Cancer control initiatives are informed by quantifying the capacity to reduce cancer burden through effective interventions. Burden measures using health administrative data are a sustainable way to support monitoring and evaluating of outcomes among patients and populations. The PREmature Mortality to IncidencE Ratio (PREMIER) is one such burden measure. We use data on Aboriginal and non-Aboriginal South Australians from 1990 to 2010 to show how PREMIER quantifies disparities in cancer burden: between populations; between sub-population cohorts where stage at diagnosis is available; and when follow-up is constrained to 24-months after diagnosis.

**Method:** PREMIER_cancer_ is the ratio of years of life expectancy lost due to cancer (YLL_cancer_) to life expectancy years at risk at time of cancer diagnosis (LYAR) for each person. The Global Burden of Disease standard life table provides referent life expectancies. PREMIER_cancer_ was estimated for the population of cancer cases diagnosed in South Australia from 1990 to 2010. Cancer stage at diagnosis was also available for cancers diagnosed in Aboriginal people and a cohort of non-Aboriginal people matched by sex, year of birth, primary cancer site and year of diagnosis.

**Results:** Cancers diagnoses (N=144,891) included 777 among Aboriginal people. Cancer burden described by PREMIER_cancer_ was higher among Aboriginal than non-Aboriginal (0.55, 95%CIs 0.52-0.59 versus 0.39, 95%CIs 0.39-0.40). Diagnoses at younger ages among Aboriginal people, 7 year higher LYAR (31.0, 95%CIs 30.0-32.0 versus 24.1, 95%CIs 24.1-24.2) and higher premature cancer mortality (YLL_cancer_**=**16.3, 95%CIs 15.1-17.5 versus YLL_cancer_**=**8.2, 95%CIs 8.2-8.3) influenced this. Disparities in cancer burden between the matched Aboriginal and non-Aboriginal cohorts manifested 24-months after diagnosis with PREMIER_cancer_ 0.44, 95%CIs 0.40-0.47 and 0.28, 95%CIs 0.25-0.31 respectively.

**Conclusion:** PREMIER described disproportionately higher cancer burden among Aboriginal people in comparisons involving: all people diagnosed with cancer; the matched cohorts; and, within groups diagnosed with same staged disease. The extent of disparities were evident 24-months after diagnosis. This is evidence of Aboriginal peoples’ substantial capacity to benefit from cancer control initiatives, particularly those leading to earlier detection and treatment of cancers. PREMIER’s use of readily available, person-level administrative records can help evaluate health care initiatives addressing this need.

## Background

Cancer is a leading cause of death and premature death globally [1, 2]. In Australia, cancer remains the largest contributor to years of life prematurely lost (YLL) despite the age standardised burden per head of population having declined by 11% from 2003 to 2011 [3]. Average burden may mask disparate trends in outcomes between and within populations [4, 5]. In the case of Aboriginal Australians (where “Aboriginal” is respectfully used to refer to people self-identifying as Aboriginal, Torres Strait Islander, or both [6]) comparable age-adjusted YLL were initially higher (52 versus 35 YLL per 1,000 population in 2003) and further increased to 55 versus 31 YLL per 1,000 population by 2013. This higher fatal burden is influenced by comparatively greater incidence of cancers with poor survival [5, 7, 8], diagnoses at more advanced stage [9-11], lower exposure to cancer treatment [9, 12], and excess case fatality concentrated in the first two-years after diagnosis [13]. Each of these influences suggest an unmet capacity to benefit from cancer control initiatives and actions including augmented cancer screening programs and addressing variations in treatment [14-16]. Such interventions need to be accompanied by relevant performance measures; measures which ensure system accountability [17], first by articulating disparity, then quantifying the capacity to benefit from prevention, early detection and intervention.

At a macro level, performance measures for population cancer outcomes [18] usually use relative survival [7, 19]. Relative survival is the ratio of observed survival among a group of people diagnosed with cancer and the expected survival of a similar, disease free group in the general population [20]. However, that method’s use can be severely limited for sub-populations of particular interest [7, 21, 22] or greatest need [22] where life tables detailing the background probabilities of death are not routinely available [23]. Such is the case with Aboriginal Australians, particularly at state and territory levels [7, 24]. An alternative is to use the Mortality to Incidence Ratio (MIR) which is the ratio of the observed cancer mortality and incidence rates in a given population in a specified time period [25, 26]. MIR is often used to illustrate disparate cancer outcomes between countries [27, 28] and the manner in which health system ranking [29] with components of cancer care such as cancer screening and treatment [28, 30-33], positively correlate with better, lower MIRs as illustrated in Figure 1 [27]. Australia’s health system is ranked thirty-second by the World Health Organization and has an average MIR of approximately 0.3, which is low by international standards and reflects well on Australia’s cancer control activities [34]. While less frequently used, MIR also describes cancer disparities within countries [35-37]. In this light, the favourable Australian average masks Aboriginal Australia’s poorer outcome of 0.5 [38].

**Figure 1.**
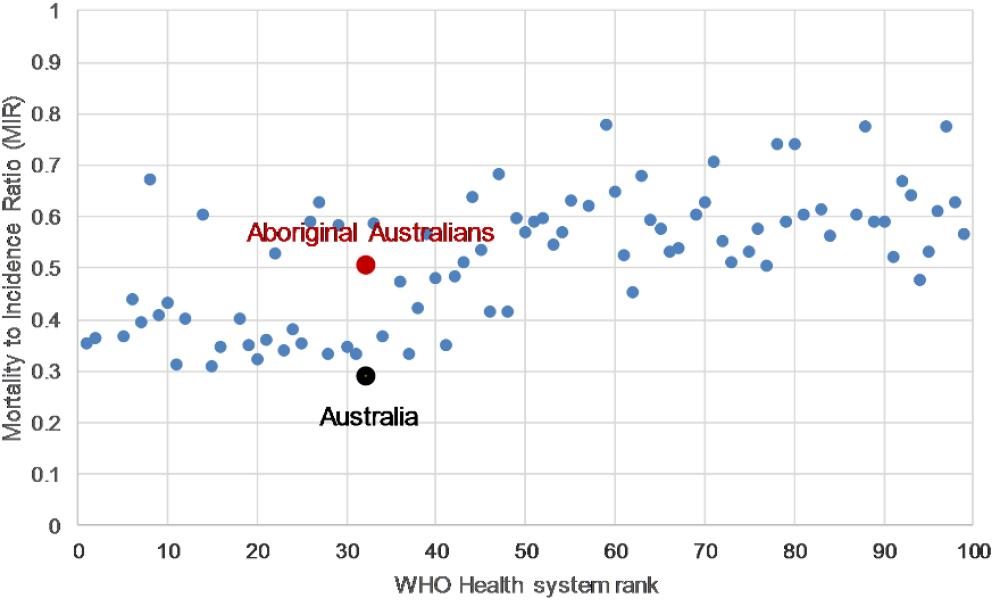
Mortality to Incidence Ratio (MIR) by the World Health Organization’s Health System ranking (Top 100)

MIR has limited application for routine performance reporting for several reasons. As with life tables [7, 21, 22], routine and/or localised estimates for calculating population incidence and mortality rates may not be readily accessible. This is the case for Aboriginal Australians with Census estimates before 2016 labelled as ‘experimental’ and yearly population updates by age and smaller geographical areas not routinely published [39]. Consequently, data availability also limits the use of MIR [40] in quantifying opportunities to tailor initiatives to the needs of relevant sub-populations [41]. In addition, population [42] and cancer registrations [5, 43] available for performance monitoring often have time lags of two years or more before their release. This is sub-optimal because disparities in cancer outcome are manifest within 24-months of diagnosis [13]. Earlier signals on outcomes are needed if we are to evaluate the effects of system change in a timely manner [44, 45].

We respond to the need to further develop performance measurement in cancer control by revising MIR with the aim of increasing comparison between and within population sub-groups and without relying on infrequently available population parameters. We do so by employing a burden of disease method and measuring the time gap [46] of optimal life expectancy [47] remaining at two critical points in a person’s experience of cancer: the age of a person’s cancer diagnosis and death from cancer. Optimal life expectancy here refers to an international standard derived from the best observed mortality rates globally [48]. By adopting this method means we re-evaluate the MIR’s underlying parameters at the person level, then aggregate results for (sub)population groups.

Consequently, we introduce the PREmature Mortality to IncidencE Ratio (PREMIER), a metric that reframes MIR within a burden of disease method. After outlining PREMIER’s components and construction, we provide four analyses demonstrating its application. *Analysis One* focuses on general disparities in cancer burden existing between populations and uses cancers diagnosed among Aboriginal and non-Aboriginal Australians. Given these populations experience differences in age and primary site of cancers diagnosed [5, 8], *Analysis Two* adjusts for those confounding variables and quantifies disparity between Aboriginal people with cancer and a cohort of cancer cases drawn from the non-Aboriginal population having the same sex, year of birth, year of cancer diagnosis and primary site. *Analysis Three* enumerates differences in PREMIER within the Aboriginal and matched non-Aboriginal cohorts on the basis of cancer stage at diagnosis. To assess the extent to which disparities in cancer burden are evident soon after diagnosis, our final *Analysis Four* evaluates cancer burden between and within the matched cohorts 24-months after diagnosis. We then consider the implications and responses to observed disparities.

## Methods

### Study design and participants

We first provide a population context of all cancer cases [excluding non-melanoma skin cancer] diagnosed among South Australians in the period 1990 to 2010 (N=144,891). A nested retrospective, matched cohort design [9, 49] is used to compare cancers cases diagnosed among Aboriginal people (N=777) with a one-to-one random selection of cancer cases among non-Aboriginals matched on the basis of sex, year of birth, primary cancer site and year of diagnosis [8]. Follow-up time is from diagnosis date to date of death, or censoring or records at 31 December 2011, whichever occurred first.

### Data sources, related measurements and definition of PREMIER

Cancer data for the South Australian population were obtained from the South Australian Cancer registry (SACR) [50] in the course of developing an advanced cancer data system within the Cancer Data and Aboriginal Disparities (CanDAD) project [51]. SACR is a population registry collating dates of International Classification of Diseases for Oncology (ICD-O-3) [52] coded diagnoses and death (attributed as cancer or non-cancer death). Specialist clinical cancer registry staff further enhanced the nested cohort study records using diagnostic and pathology records available to SACR to include cancer stage at diagnosis using Surveillance, Epidemiology, and End Results Program methodologies [53]. Stage at diagnosis categories included: *localised -* confined to tissue of origin; *regional -* invaded adjacent tissue or regional nodes; *distant/unknown -* spread to distant lymph nodes or other organ sites; leukaemia; or insufficient staging data were available.

MIR parameters of mortality and incidence are reframed within a burden of disease framework in the following manner. Mortality among cancer cases is quantified using YLL [54, 55], the amount of life expectancy remaining at time at which death attributed to cancer occurred. Incidence is quantified using expected Life Years at Risk (LYAR) [56], that is, the amount of life expectancy remaining at time at which cancer diagnosis occurred. Both YLL and LYAR represent the years of optimal life expectancy remaining at the age a given event occurs. That optimal life expectancy, which is subsequently used as a standard against which other measures are made, was previously derived for the global burden of disease study using the lowest age-specific risk of death observed in populations greater than 5 million individuals across the world (further details are available in Appendix Table 18, p503 [54]). In the case of YLL, the relevant event is the age at death while LYAR refers to age at diagnosis.

We make three assumptions in adopting those standard life expectancy estimates. First, we assume it is fair that all people aspire to optimal life expectancy because health differentials between sub-populations are influenced through societal and environmental risk factor exposures [47, 48] rather than fixed biological determinants aside from age. Second, we assume a uniform estimate of life expectancy across time, place and circumstance facilitates fair comparisons, regardless of changing geographic or sub-population specific mortality rates. We also assume a consistent method to deriving measures facilitates comparison between those measures and such comparisons are valuable.

PREMIER represents the amount of life expectancy lost as a fraction of life expectancy remaining at the time a sentinel health event is diagnosed. In the case of premature loss of life from cancer death after cancer diagnosis (PREMIER_cancer_), this is the ratio of years of life lost attributed to cancer (YLL_cancer_) to expected life years at risk at the time of cancer diagnosis (LYAR) represented as:

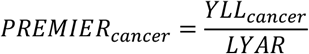

As a fraction of YLL and LYAR, PREMIER ranges from 0, where death after cancer diagnosis does not occur within the observation period, to 1, where death occurs at the same age as diagnosis. As an example, a person diagnosed with cancer at age 55 is taken as having 32.9 years of life expectancy remaining, thus LYAR is 32.9. Where death from cancer follows at age 65 the remaining life expectancy represents 23.8 years of life lost to cancer, YLL_cancer_. PREMIER_cancer_ is 23.8 / 32.9, or 0.72, indicating that 72% of life expectancy at time of diagnosis was subsequently lost.

Individual PREMIER, and its LYAR and YLL components, can be grouped across population groups, or cohorts of people diagnosed with cancer. PREMIER can refer to a variety of observation periods. For instance, populations or cohorts may be observed for: varying periods from time of diagnosis to right-censoring of observations at a given date; a fixed period after cancer diagnosis; or, a combination of the two.

### Statistical analysis

Under the heading of *Risk*, we summarise the mean age at cancer diagnosis and the accompanying LYAR. Subsequent *Loss* to premature mortality describes the number and mean age of deaths observed and attributed to cancer by SACR. Where deaths were not attributed to cancer, YLL_cancer_ is zero. The *Loss to Risk ratio*, comprises the averaged PREMIER_cancer_ for individuals within each group.

Table 1 includes three groups of cancer cases: the population of cancer cases diagnosed from 1990 to 2010 among non-Aboriginal South Australians; cancer cases diagnosed among Aboriginal South Australians in the same period; and, a matched cohort of cancer cases among non-Aboriginal people. Table 2 focuses on the Aboriginal and non-Aboriginal cohorts disaggregated by stage at diagnosis. Table 3 repeats this focus while limiting observation time to a maximum of 24-months after diagnosis.

**Table 1:** Cancer diagnoses, premature mortality and PREMIER_cancer_, South Australia 1990-2010*.

|  | Cases among non-Aboriginal |  |  |  | Cases among Aboriginal |  |  |  | Matched cases among non-Aboriginal<sup>#</sup> |  |  |  |
| --- | --- | --- | --- | --- | --- | --- | --- | --- | --- | --- | --- | --- |
|  | N | % | Mean | 95% CIs | N | % | Mean | 95% CIs | N | % | Mean | 95% CIs |
| **Risk** |  |  |  |  |  |  |  |  |  |  |  |  |
| Age at diagnosis (years) | 144,114 | 100.0% | 65.5 | 65.4-65.6 | 777 | 100.0% | 57.7 | 56.6-58.8 | 777 | 100.0% | 58.5 | 57.4-59.5 |
| Life Years at Risk (LYAR) |  |  | 24.1 | 24.1-24.2 |  |  | 31.0 | 30.0-32.0 |  |  | 30.3 | 29.3-31.3 |
| **Loss** |  |  |  |  |  |  |  |  |  |  |  |  |
| Cancer deaths* and age (years) | 62,936 | 43.7% | 71.7 | 71.6-71.8 | 461 | 59.3% | 61.5 | 60.2-62.9 | 340 | 43.8% | 63.7 | 62.1-65.2 |
| Years of life lost from cancer (YLL<sub>cancer</sub>) |  |  | 8.2 | 8.2-8.3 |  |  | 16.3 | 15.1-17.5 |  |  | 11.2 | 10.1-12.3 |
| **Loss:Risk ratio** |  |  |  |  |  |  |  |  |  |  |  |  |
| Premature mortality to incidence ratio (PREMIER<sub>cancer</sub>) |  |  | 0.39 | 0.39-0.40 |  |  | 0.55 | 0.52-0.59 |  |  | 0.40 | 0.37-0.44 |

\*Among observations right-censored at 31/12/2011
<sup>#</sup> Randomly selected cancer cases among non-Aboriginal people matched one to one with cases among Aboriginal by sex, year of birth, year of diagnosis and primary cancer site

**Table 2:** Cancer diagnoses, premature mortality and PREMIER_cancer_ by stage at diagnosis, South Australia 1990-2010*.

|  | Localised at diagnosis |  |  |  |  |  |  |  | Regional spread at diagnosis |  |  |  |  |  |  |  | Distant/Unknown spread at diagnosis |  |  |  |  |  |  |  |
| --- | --- | --- | --- | --- | --- | --- | --- | --- | --- | --- | --- | --- | --- | --- | --- | --- | --- | --- | --- | --- | --- | --- | --- | --- |
|  | Aboriginal |  |  |  | Matched non-Aboriginal<sup>#</sup> |  |  |  | Aboriginal |  |  |  | Matched non-Aboriginal<sup>#</sup> |  |  |  | Aboriginal |  |  |  | Matched non-Aboriginal<sup>#</sup> |  |  |  |
|  | N | % | Mean | 95%Cls | N | % | Mean | 95%Cls | N | % | Mean | 95%Cls | N | % | Mean | 95%Cls | N | % | Mean | 95%Cls | N | % | Mean | 95%Cls |
| Risk |  |  |  |  |  |  |  |  |  |  |  |  |  |  |  |  |  |  |  |  |  |  |  |  |
| Age at diagnosis | 289 | 100.0% | 58.4 | 56.5-60.3 | 390 | 100.0% | 57.8 | 56.2-59.3 | 155 | 100.0% | 55.5 | 53.2-57.8 | 132 | 100.0% | 57.9 | 55.4-60.5 | 333 | 100.0% | 58.1 | 56.4-59.8 | 255 | 100.0% | 59.8 | 57.9-61.7 |
| LYAR |  |  | 30.4 | 28.7-32.2 |  |  | 30.9 | 29.5-32.4 |  |  | 32.8 | 30.7-34.9 |  |  | 30.7 | 28.4-33.0 |  |  | 30.6 | 29.1-32.1 |  |  | 29.1 | 27.4-30.8 |
| Loss |  |  |  |  |  |  |  |  |  |  |  |  |  |  |  |  |  |  |  |  |  |  |  |  |
| Cancer deaths* | 101 | 34.9% | 65.9 | 63.0-68.9 | 100 | 25.6% | 64.9 | 61.6-68.2 | 93 | 60.0% | 58.9 | 56.1-61.7 | 59 | 44.7% | 63.1 | 59.6-66.7 | 267 | 80.2% | 60.8 | 59.1-62.6 | 181 | 71.0% | 63.2 | 61.1-65.2 |
| YLL<sub>cancer</sub> |  |  | 8.3 | 6.7-9.9 |  |  | 6.3 | 5.0-7.6 |  |  | 17.8 | 15.1-20.6 |  |  | 11.6 | 9.0-14.2 |  |  | 22.5 | 20.8-24.3 |  |  | 18.5 | 16.6-20.4 |
| Loss:Risk ratio |  |  |  |  |  |  |  |  |  |  |  |  |  |  |  |  |  |  |  |  |  |  |  |  |
| PREMIER<sub>cancer</sub> |  |  | 0.30 | 0.25-0.35 |  |  | 0.22 | 0.18-0.26 |  |  | 0.56 | 0.49-0.64 |  |  | 0.41 | 0.33-0.49 |  |  | 0.77 | 0.73-0.81 |  |  | 0.68 | 0.62-0.73 |

\*Among observations right-censored at 31/12/2011
<sup>#</sup> Randomly selected cancer cases among non-Aboriginal people matched one to one with cases among Aboriginal by sex, year of birth, year of diagnosis and primary cancer site

**Table 3:** Cancer diagnoses, premature mortality and PREMIER_cancer_ at 24-months by stage at diagnosis, South Australia 1990-2010*.

|  |  | All cancers |  |  |  |  |  |  |  | Localised at diagnosis |  |  |  |  |  |  |  |
| --- | --- | --- | --- | --- | --- | --- | --- | --- | --- | --- | --- | --- | --- | --- | --- | --- | --- |
|  |  | Aboriginal |  |  |  | Matched non-Aboriginal<sup>#</sup> |  |  |  | Aboriginal |  |  |  | Matched non-Aboriginal<sup>#</sup> |  |  |  |
|  |  | N | % | Mean | 95%Cls | N | % | Mean | 95%Cls | N | % | Mean | 95%Cls | N | % | Mean | 95%Cls |
| Risk | Age at diagnosis | 777 | 100.0% | 57.7 | 56.6-58.8 | 777 | 100.0% | 58.5 | 57.4-59.5 | 289 | 100.0% | 58.4 | 56.5-60.3 | 390 | 100.0% | 57.8 | 56.2-59.3 |
|  | LYAR |  |  | 31.0 | 30.0-32.0 |  |  | 30.3 | 29.3-31.3 |  |  | 30.4 | 28.7-32.2 |  |  | 30.9 | 29.5-32.4 |
| Loss | Cancer deaths<sub>24-months</sub>* |  |  |  |  |  |  |  |  |  |  |  |  |  |  |  |  |
|  | Age at death<sub>cancer 24-months</sub> | 346 | 44.5% | 60.4 | 58.9-61.9 | 224 | 28.8% | 63.5 | 61.6-65.4 | 51 | 17.6% | 63.0 | 58.5-67.5 | 40 | 10.3% | 64.4 | 59.3-69.4 |
|  | YLL<sub>cancer 24-months</sub> |  |  | 12.7 | 11.5-13.9 |  |  | 7.4 | 6.5-8.4 |  |  | 4.7 | 3.3-6.0 |  |  | 2.6 | 1.7-3.5 |
| Loss:Risk ratio |  |  |  |  |  |  |  |  |  |  |  |  |  |  |  |  |  |
| PREMIER<sub>cancer 24-months</sub> |  |  |  | 0.44 | 0.40-0.47 |  |  | 0.28 | 0.25-0.31 |  |  | 0.17 | 0.13-0.21 |  |  | 0.10 | 0.07-0.13 |

|  |  | Regional spread at diagnosis |  |  |  |  |  |  |  | Distant/Unknown spread at diagnosis |  |  |  |  |  |  |  |
| --- | --- | --- | --- | --- | --- | --- | --- | --- | --- | --- | --- | --- | --- | --- | --- | --- | --- |
|  |  | Aboriginal |  |  |  | Matched non-Aboriginal<sup>#</sup> |  |  |  | Aboriginal |  |  |  | Matched non-Aboriginal<sup>#</sup> |  |  |  |
|  |  | N | % | Mean | 95%Cls | N | % | Mean | 95%Cls | N | % | Mean | 95%Cls | N | % | Mean | 95%Cls |
| Risk | Age at diagnosis | 155 | 100.0% | 55.5 | 53.2-57.8 | 132 | 100.0% | 57.9 | 55.4-60.5 | 333 | 100.0% | 58.1 | 56.4-59.8 | 255 | 100.0% | 59.8 | 57.9-61.7 |
|  | LYAR |  |  | 32.8 | 30.7-34.9 |  |  | 30.7 | 28.4-33.0 |  |  | 30.6 | 29.1-32.1 |  |  | 29.1 | 27.4-30.8 |
| Loss | Cancer deaths<sub>24-months</sub>* |  |  |  |  |  |  |  |  |  |  |  |  |  |  |  |  |
|  | Age at death<sub>cancer 24-months</sub> | 67 | 43.2% | 58.5 | 55.2-61.8 | 37 | 28.0% | 63.4 | 58.4-68.3 | 228 | 68.5% | 60.4 | 58.5-62.3 | 147 | 57.6% | 63.3 | 61.0-65.6 |
|  | YLL<sub>cancer 24-months</sub> |  |  | 13.0 | 10.3-15.7 |  |  | 7.2 | 4.9-9.6 |  |  | 19.5 | 17.7-21.4 |  |  | 15.0 | 13.0-16.9 |
| Loss:Risk ratio |  |  |  |  |  |  |  |  |  |  |  |  |  |  |  |  |  |
| PREMIER<sub>cancer 24-months</sub> |  |  |  | 0.42 | 0.35-0.50 |  |  | 0.27 | 0.20-0.35 |  |  | 0.68 | 0.63-0.72 |  |  | 0.57 | 0.51-0.63 |

\*Among observations right-censored at a maximum of 24 months after diagnosis or 31/12/2011
<sup>#</sup> Randomly selected cancer cases among non-Aboriginal people matched one to one with cases among Aboriginal by sex, year of birth, year of diagnosis and primary cancer site

Our multivariable analysis used the matched cohorts to evaluate the relationship between: PREMIER_cancer_ at 24-months after diagnosis (PREMIER_cancer 24-months_) as the outcome with Aboriginality as the exposure and, cancer stage at diagnosis as a covariate. Interactions between Aboriginality and stage at diagnosis were also examined. We used fractional response regression [57], a quasi-likelihood estimation method available within Stata 15.1 as *fracreg* [58], and assumed a probit model for the conditional mean. This approach accommodates PREMIER’s attributes as: a fraction of two continuous quantities with life expectancy lost as numerator, life expectancy at time of diagnosis as denominator; having a denominator which is also the maximum value for the numerator; and, thus having values in the range of 0 to 1 inclusive. We clustered the data by the cohorts’ matched pairs and report 95% confidence intervals (95% CIs) based on robust standard errors. We report the modelled parameter coefficients which provide the sign of each covariate’s effect on PREMIER_cancer 24-months_. However, because the coefficients are difficult to interpret we also assessed the simultaneous average marginal effects of Aboriginality and stage at diagnosis on the proportion of life at risk lost in the 24-month period from diagnosis. That is, we report the change in PREMIER_cancer 24-months_ where the cancer case involved an Aboriginal person rather than non-Aboriginal; and localised or distant stages rather than regional stage disease at diagnosis.

## Results

### Cancer burden between population groups

Table 1 shows SACR recorded 144,891 invasive cancer diagnoses among South Australians from 1990 to 2010. Cancer diagnoses among Aboriginal people accounted for a small number of those cases (N=777) and these are described in detail elsewhere [8]. Notably though, the latter cases were diagnosed at considerably younger age (57.7 years) compared to those among non-Aboriginal people (65.5 years). Consequently, life expectancy at risk at time of cancer diagnosis was almost 7 years higher among Aboriginal people with LYAR=31.0 (95% CIs 30.0-32.0) compared to the non-Aboriginal average of LYAR=24.1 (95%CIs 24.1-24.2). Proportionately more case fatalities, and at younger average age, were also observed among Aboriginal people with cancer. Taken together, average loss to premature mortality from cancer among Aboriginal cases was twice that of the broader group of non-Aboriginal cases (YLL_cancer_**=**16.3, 95%CIs 15.1-17.5 versus YLL_cancer_**=**8.2, 95%CIs 8.2-8.3). In turn, PREMIER_cancer_ was markedly higher among Aboriginal compared to non-Aboriginal cases at 0.55 (95%CIs 0.52-0.59) versus 0.39 (95%CIs 0.39-0.40) respectively.

### Cancer burden between and within matched cohorts

Table 1 also compares cases among Aboriginal people compared to a randomly selected cohort of diagnoses among non-Aboriginal cases (N=777) matched by sex, year of birth, year of diagnosis and primary cancer site. LYAR among the Aboriginal and non-Aboriginal cohort are therefore equivalent because of age matching. Fewer case fatalities at comparatively older ages among the non-Aboriginal cohort led to an average YLL_cancer_ at 11.2 (95% CIs 10.1-12.3) and PREMIER_cancer_ at 0.40 (95% CIs 0.37-0.44) which were markedly lower than their matched Aboriginal contemporaries with PREMIER_cancer_=0.55 (95%CIs 0.52-0.59). Indeed, PREMIER_cancer_ for all non-Aboriginal and the subset of cases within the non-Aboriginal cohort were very similar (0.39, 95%CIs 0.39-0.40 and 0.40, 95% CIs 0.37-0.44 respectively).

Table 2 disaggregates Aboriginal and matched non-Aboriginal cohort results by stage at diagnosis. Cancers among Aboriginal people were more likely to involve distantly spread disease (n=333 or 42.8% of cases) than among non-Aboriginal people (n=255 or 32.8% of cases). Within each stage at diagnosis cancer case fatality was relatively more common among Aboriginal than non-Aboriginal people. Also, the average age at cancer death was lower among Aboriginal people than non-Aboriginal people diagnosed with regionally staged disease (58.9 versus 63.1 years) and distant staged disease (60.8 versus 63.2 years). Both factors contributed to markedly greater average YLL_cancer_ in the Aboriginal cohort than the non-Aboriginal cohort with differences ranging from 2.0 (95%CIs 1.7-2.3) in localised stage to 6.2 (6.1-6.2) in regionally spread disease. For both cohorts, PREMIER_cancer_ increased as cancer spread at diagnosis increased. However, PREMIER_cancer_ also showed the relative amount of life at risk and subsequently lost was higher within the Aboriginal cohort at each stage of disease at diagnosis.

### Cancer burden two years after diagnosis

Table 3 shows cohort outcomes up to two years after cancer diagnosis. Case fatality increased as stage at diagnosis increased from local to regional to distant stages with consistently higher loss observed among Aboriginal compared to non-Aboriginal people. Again, age at cancer death was younger among Aboriginal people than non-Aboriginal people for each stage at diagnosis. Average YLL_cancer_ was also higher among Aboriginal cases at each stage of disease at diagnosis. Consequently, PREMIER_cancer_ differed between cohorts 24-months after diagnosis with higher losses among Aboriginal (PREMIER_cancer 24-months_=0.44, 95%CIs 0.40-0.47) than non-Aboriginal (PREMIER_cancer 24-months_=0.28, 95%CIs 0.25-0.31). This difference of 0.16 in the limited 24-month follow-up period (using PREMIER_cancer 24-months_) was very similar to the difference of 0.15 observed across the full observation period (using PREMIER_cancer_).

PREMIER_cancer 24-months_ also differed within cohorts and increased as stage at diagnosis increased. For example, point estimates for PREMIER_cancer 24-months_ within the Aboriginal cohort increased from 0.17 in cases of localised disease to 0.68 where disease spread was distant or unknown, an overall change of 0.51. Overall change within the non-Aboriginal cohort was slightly less at 0.47 and ranged from 0.10 in localised disease to 0.57 in distant spread disease.

### Multivariable analysis

Table 4 shows the association between life at risk and life subsequently lost up to 24-months after cancer diagnosis in the cohorts and the concurrent effects of Aboriginality and stage at diagnosis. Both Aboriginality and advancing disease stage at diagnosis were associated with higher PREMIER_cancer_. The model’s marginal effects indicate Aboriginal cases experienced an average of 0.10 or 10% (95%CIs 0.06-0.14) higher PREMIER_cancer_ than non-Aboriginal cohort cases diagnosed with the same stage of disease. Simultaneously, and when compared to regionally spread disease at diagnosis, localised disease was associated with 0.21 or 21% (95%CIs 0.14-0.27) lower PREMIER_cancer_ and distant/unknown spread with 0.27 or 27% (95%CIs 0.20-0.34) higher PREMIER_cancer_. No further interaction of the effects of Aboriginality by stage at diagnosis was evident.

**Table 4:** Fractional outcome regression and average marginal effects on PREMIER_cancer_ at 24-months, South Australia 1990-2010*.

|  |  | Model for PREMIER<sub>cancer 24-months</sub> |  |  |  | Average marginal effects<sup>#</sup> |  |  |  |
| --- | --- | --- | --- | --- | --- | --- | --- | --- | --- |
|  |  | Coef. | 95% CIs | z | p>\|z\| | dy/dx | 95% CIs | z | p>\|z\| |
| Aboriginal | No | 0.00 | Reference |  |  | 0.00 | Reference |  |  |
|  | Yes | 0.33 | 0.21-0.45 | 5.46 | <0.001 | 0.10 | 0.06-0.14 | 5.43 | <0.001 |
| Stage at diagnosis |  |  |  |  |  |  |  |  |  |
|  | Localised | -0.72 | -0.92--0.53 | -7.38 | <0.001 | -0.21 | -0.27--0.14 | -6.92 | <0.001 |
|  | Regional | 0.00 | Reference |  |  | 0.00 | Reference |  |  |
|  | Distant/unknown | 0.70 | 0.52-0.89 | 7.48 | <0.001 | 0.27 | 0.20-0.34 | 7.81 | <0.001 |
| Constant |  | -0.56 | 0.30-0.52 | -6.67 | <0.001 |  |  |  |  |

\* Using a randomly selected cancer cases among non-Aboriginal people matched one to one with cases among Aboriginal by sex, year of birth, year of diagnosis and primary cancer site with observations right censored at a maximum of 24 months after diagnosis or at 31/12/2011
<sup>#</sup> Average marginal effects represent the change in PREMIER<sub>cancer 24-months</sub>, the outcome variable, when moving from a predictor variable's reference category

## Discussion

PREMIER combines life expectancy at the time of cancer diagnosis and the resultant loss of life due to cancer death in order to quantify cancer burden. This is calculated for each person diagnosed with subsequent aggregation to groups. Our first analysis demonstrated PREMIER’s application in describing disparities in cancer burden for the entire population of invasive cancers diagnosed among South Australians. PREMIER described substantially higher cancer burden among the population of Aboriginal people with cancer compared to other South Australians (PREMIER_cancer_ of 0.55 versus 0.39). These differences were bought about by Aboriginal South Australians with cancer having lower average age and more life expectancy (7 years) at risk of loss while also experiencing higher average premature mortality loss due to higher case fatality (59.3% versus 43.7%) and younger age at death (62 versus 72 years). Our second analysis focussed on Aboriginal and non-Aboriginal cohorts with equivalent sex, age, year of diagnosis and primary cancer site. While life expectancy at diagnosis was equivalent, PREMIER enumerated 15% more cancer burden among Aboriginal South Australians with cancer (PREMIER_cancer_ of 0.55 versus 0.40). This was influenced by more frequent cancer deaths (59.3% versus 43.8%) and these deaths being at a younger age (61.5 versus 63.7 years). With the availability of stage at diagnosis for the cohorts, we then considered the variation of cancer burden within the cohorts. In both cohorts PREMIER increased as stage increased from local to regional to distant spread. In addition, PREMIER remained higher among Aboriginal people at each stage (PREMIER_cancer_=0.30 versus 0.22 for localised disease; 0.56 versus 0.41 for regional spread; and, 0.77 versus 0.68 for distant spread). These disparities by stage and Aboriginality were not only apparent for the broader observation period. They were fully manifested 24-months after diagnosis and our fourth analysis showed 16% higher cancer burden among Aboriginal than non-Aboriginal contemporaries (PREMIER_cancer 24-months_ of 0.44 versus 0.28 respectively). Disparity of this size then continued across longer term observations.

Our analyses align with other reports of MIR, the ratio of observed cancer mortality and incidence rates in a given population in a specified time period, which describe intra-country disparities in cancer outcomes. For example, MIR differences between Black (MIR=0.48) and White (MIR=0.40) in South Carolina are clear [35, 37], yet recent differences between Aboriginal (MIR=0.51) and Australia generally (MIR=0.30) are even more pronounced [38]. These disparate results are echoed by PREMIER within the population of South Australians diagnosed with cancer where substantially more cancer burden among Aboriginal than non-Aboriginal (PREMIER_cancer_=0.55 versus 0.39 respectively) was quantified.

There are notable points of difference between MIR and PREMIER though. MIR makes use of mortality and incidence rates calculated on people diagnosed or dying in any given period. Those dying may have been diagnosed in different time periods meaning different groups of people are being compared [19]. One of the consequences of this back-scattering of incident cases is to make it difficult to observe rapid changes in prognosis [19]. PREMIER however, draws directly on each individual case for both numerator (LYAR) and denominator (YLL). Because incidence and mortality are observed within the same person the need to adjust for back-scattering is avoided. This is an advantage because it enables PREMIER to provide an earlier signal on cancer outcomes. Earlier measures can inform timely evaluations of system change, particularly system change aimed at improving outcomes within 24-months of diagnosis, a time when disparities are entrenched but also able to be detected using PREMIER.

PREMIER’s perspective on cancer burden is relevant to evidence-based policy development in cancer control [59] in other ways. For example, PREMIER’s estimation provides absolute measures of life at risk and life lost from cancer in a manner that is useful to planning activities. This is achieved by anchoring age at diagnosis and age at cancer death against a defined, optimal outcome. By describing disparities in age at diagnosis LYAR determined the amount of life expectancy amenable to change by preventing cancer, or at least deferring cancer incidence to later ages, through reduced exposure to cancer risks. As a relative measure, PREMIER revealed disparities across stage at diagnosis where more advanced disease led to higher cancer mortality and higher PREMIER. This information can help prioritise activities leading to earlier case detection and increased participation in cancer screening activities to detect cancers at an earlier stage. PREMIER also demonstrated an ability to enumerate disparities in cancer burden associated with stage and ethnicity 24-months after cancer diagnosis, a time during which people are more likely to be receiving care through health services [45]. This becomes particularly useful in supporting activities that promote access [60], uptake and quality [15, 61] of effective and available cancer treatments. In short, PREMIER enumerates people’s capacity to benefit from cancer control initiatives involving prevention, early detection and treatment and thus contributes to prioritising health system activities.

Similarly, while we report aggregated outcomes, it is important to remember PREMIER is calculated for each individually diagnosed case which become available for grouping and analysed in many configurations. We grouped observations by Aboriginality, however groups could be based on: shared area level geography; socio-economic position; or, by attending a certain service or receiving the care of particular provider. This adaptability is not only relevant to policy and planning but has further application in relating system performance to outcomes for individuals and the population groups to whom they belong [41]. PREMIER offers a robust and contemporary measure of performance with which to assess the effectiveness of early detection and treatment efforts. This is because PREMIER is free of the immediate need for background population information and time lags in reporting are reduced with counting and observations beginning as soon as diagnosis is made. This suggests the use of clinical records for reporting at patient (micro) and service (meso) levels in the first instance. As the underlying cancer and mortality records are integrated into population registries as we have used, macro level reporting for populations and the whole of system can follow. Information at these varying levels lend themselves to continued quality improvement processes and ongoing applied research. The use of existing, routine administrative data also helps address the evaluation needs of health services and government [62] while promoting public accountability [63]. Indeed, incorporating YLL within PREMIER facilitates comparison with other health system indicators and targets around reducing avoidable and premature mortality, particularly among vulnerable populations [63].

PREMIER has other strengths. Our analyses demonstrate the feasibility of assessing PREMIER using existing, routine, administrative and/or clinical records which also suggests it is readily sustainable. Other parameters from hospital systems could inform stratification within patient groups, for example, by stage at diagnosis. As cancer mortality outcomes improve and it becomes increasingly important to assess patient morbidity, the burden of disease method also provides for health adjusting the age relevant life expectancy and incorporating this into PREMIER estimates [56, 64]. In the meantime, PREMIER responds to the call for ever-increasing comparability and granularity in reporting [64] in two ways. We showed PREMIER’s comparability across populations and within small cohort groups. Further comparison with the wider Australian community, or even globally and for other time periods is quite possible because by measuring against the same, global standard. PREMIER has additional scope to generalise across conditions such as stroke or heart attack where there are definitive times of diagnosis enabling assessment of LYAR and subsequent YLL components. This would inform further comparison between and within people groups on the basis of health condition.

### Limitations

PREMIER has several limitations. Interpreting relative outcome measures expressed as ratios which depend on different numerators and denominators is challenging. It is also a commonly occurring issue when considering issues of health disparity [65]. Our suggested response is to accompany PREMIER with reports of LYAR and YLL as absolute measures based on life expectancy. This raises the major limitation of PREMIER in that both LYAR and YLL are predicated on a global standard life table while local life expectancy for population groups of interest will likely be different. That is, PREMIER makes use of two biased measures and overestimates outcome disparities [66, 67] suggesting a prudent approach to its use as recommended with other survival methods [68]. The counter argument is to avoid bias by using population specific life tables [69-71]. However, life tables reflecting jurisdiction or group averages do not necessarily remedy the issue because such averages may mask considerable variation within the relevant jurisdictions or population group. For example, average life expectancy within one US county having the benefit of one of the highest observed life expectancies at birth was recently shown to subsume variations of up to 18 years among males and 15 years for females [72]. Nevertheless, when relevant life tables become available, the bias within our analysis can be approximated as done in other instances assessing the need for intra-country socio-economic position life tables [68]. Until such time though, our analysis makes use of the fall-back recommendation of using cancer specific mortality. This is justified because where health inequities exist, it is unacceptable to wait until complete information is to hand before acting. Therefore, we adopt an imperfect but well based and transparent method to quantifying health inequity by measuring against a gold standard, optimal outcome. In our case, this outcome is a standard attained by some but markedly less so by others within the same country and served by the same universal, healthcare system.

We further acknowledge our analysis of PREMIER did not account for the influence of comorbid conditions [73, 74]. In their own right, these are a major point of difference in the health status of Aboriginal and other Australians. However, PREMIER estimates for all-causes of death among people with cancer are easily calculated. Where higher risk of death from non-cancer causes are experienced [23] PREMIER estimates would increase and potentially exacerbate the disparities we documented. Other cancer survival studies do in fact report changes in the risk of death from cancer or non-cancer causes in the five years after cancer diagnosis [23] and this issue will benefit from further investigation.

## Conclusion

We demonstrated PREMIER’s application in quantifying cancer burden disparities using Aboriginal and non-Aboriginal comparisons in South Australia. Cancer burden was markedly higher among Aboriginal people than non-Aboriginal in all comparisons based on: all people diagnosed with cancer; groups matched by sex, age, primary site and year of diagnosis; and, within groups experiencing similarly staged disease at diagnosis. Importantly, the extent of disparities were evident 24-months after diagnosis and persisted at similar levels thereafter. This points to a substantial capacity to benefit from improved cancer control initiatives among Aboriginal people, particularly those health system activities aimed at earlier detection and treatment of cancers. Our analyses also suggest PREMIER’s use of readily available, person-level information can provide important information helping evaluate person-centred cancer care as one dimension of high-quality health care delivery addressing this need.

## Data Availability

The datasets generated and/or analysed during the current study are not publicly available due to privacy reasons, including the provisions of the Australian Privacy Principles. The study's data comprised of de-identified unit record administrative records and were used under privileged arrangements set out in a study specific confidentiality deed. The data cannot be accessed by another party without relevant data custodian and human research ethics approvals.

## Abbreviations

95% CIs: 95% confidence intervals
CanDAD: Cancer Data and Aboriginal Disparities
LYAR: Life Years at Risk
PREMIER: PREmature Mortality to IncidencE Ratio calculated as YLL/LYAR for each case
PREMIER_cancer_: PREmature Mortality to IncidencE Ratio calculated as YLL_cancer_/LYAR for each case
PREMIER_cancer 24-months_: PREmature Mortality to IncidencE Ratio up to 24-months after diagnosis calculated as YLL_cancer 24-months_/LYAR for each case
PROMs: Patient Reported Outcome Measures
YLL: Years of Life Lost
YLL_cancer_: Years of Life Lost associated with cancer death
YLL_cancer 24-months_: Years of Life Lost associated with cancer death up to 24-months after diagnosis
SACR: South Australian Cancer Registry

## Declarations

### Ethics approval and consent to participate

South Australia’s Aboriginal Health Research Ethics Committee (AHREC 04-12-461) and SA Health’s Human Research Ethics Committee (SA Health HREC HREC/12/SAH/35) approved the use of population cancer registry records. CanDAD’s Aboriginal Community Reference Group governance ensured alignment of the study protocol with South Australian Aboriginal Health Research Accord principles [75].

### Consent for publication

Not applicable.

### Authors’ contributions

DB conceived the project, performed the analyses and drafted the manuscript; JL, JK, AB and DR made important contributions to operationalising this study, interpreting the statistical analysis, and revised the manuscript. All authors read and approved the final version of the manuscript.

## Acknowledgements

Dr Graeme Tucker for statistical advice and the Cancer Data and Aboriginal Disparities (CanDAD) Aboriginal Community Reference Group (ACoRG), particularly Aunty Roslyn Weetra.

